# Understanding the value of clinical symptoms of COVID-19. A logistic regression model

**DOI:** 10.1101/2020.10.07.20207019

**Authors:** Pedro E. Fleitas, Jorge A. Paz, Mario I. Simoy, Carlos Vargas, Rubén O. Cimino, Alejandro J. Krolewiecki, Juan P. Aparicio

**Affiliations:** Instituto de Investigaciones de Enfermedades Tropicales (IIET-CONICET), Universidad Nacional de Salta. San Ramón de la Nueva Orán, Salta, Argentina; Cátedra de Química Biología, Facultad de Ciencias Naturales, Universidad Nacional de Salta. Salta, Salta, Argentina; Instituto de Estudios Laborales y del Desarrollo Económico (IELDE), Consejo Nacional de Investigaciones Científicas y Técnicas (CONICET), Universidad Nacional de Salta. Salta, Salta, Argentina; Instituto de Investigaciones en Energía no Convencional (INENCO), Consejo Nacional de Investigaciones Científicas y Técnicas (CONICET), Universidad Nacional de Salta, Salta, Argentina; Instituto Multidisciplinario sobre Ecosistemas y Desarrollo Sustentable, Universidad Nacional del Centro de la Provincia de Buenos Aires. Tandil, Buenos Aires, Argentina; Simon A. Levin Mathematical, Computational and Modeling Sciences Center, Arizona State University, Tempe, USA

## Abstract

**Background:** The new coronavirus SARS-CoV-2, the causative agent of COVID-19, is responsible for the current pandemic outbreak worldwide. However, there is limited information regarding the set of specific symptoms of COVID-19. Therefore, the objective of this study was to describe the main symptoms associated with COVID-19 to aid in the clinical diagnosis for the rapid identification of cases.

**Methods and findings:** A cross sectional study of all those diagnosed by RT-PCR for SARS-CoV-2 between April 1 and May 24 in Argentina was conducted. The data includes clinical and demographic information from all subjects at the time of presentation, which were uploaded to the centralized national reporting system at health centers. A total of 67318 individuals were included in this study, where 12% tested positive for SARS-CoV-2. The study population was divided in two age groups, a group aged 0 to 55 years-old (<56 group), (median = 32, n=48748) and another group aged 56 to 103 years-old (≥56 group) (median =72, n=18570). Multivariate logistic regression analyses showed that out of a total of 23 symptoms, only five were found to have a positive association with COVID-19: anosmia (odds ratio [OR] 10.40, 95% confidence interval [CI] 8.20-13.10, <56 group; OR 6.09 CI 3.05-12.20, ≥56 group) dysgeusia (OR 3.67, CI 2.7-4.9, <56 group; OR 3.53 CI 1.52-8.18, ≥56 group), low grade fever (37.5-37.9° C) (OR 1.61, CI 1.20-2.05, <56 group; OR 1.80 CI 1.07-3.06, ≥56 group), cough (OR 1.20, CI 1.05-1.38, <56 group; OR 1.37 CI 1.04-1.80, ≥56 group) and headache only in <56 group (OR 1.71, CI 1.48-1.99). In turn, at the time of presentation, the symptoms associated with respiratory problems: chest pain, tachypnea, dyspnea, respiratory failure and use of accessory muscles for breathing, had a negative association with COVID-19 (OR <1) or did not present statistical relevance (OR = 1).

With the intention of helping the clinical diagnosis, we designed a model to be able to identify possible cases of COVID-19. This model included 16 symptoms, the age and sex of the individuals, and was able to detect 80% of those infected with SARS-CoV-2 with a specificity of 46%.

**Conclusions:** The analysis of symptoms opens the opportunity for a guidance and improved symptoms based definition of suspected cases of COVID-19, where multiple factors (age, sex, symptoms and interaction between symptoms) are considered. We present a tool to help identify COVID-19 cases to provide quick information to aid decision-making by health personnel and program managers.

## Introduction

The new coronavirus SARS-CoV-2, the causative agent of COVID-19, is responsible for the current pandemic outbreak as declared by the World Health Organization on January 30th 2020 (1). By September 2020, the number of people infected with SARS-CoV-2 is over 28 million worldwide with more than 900 thousand deaths (2). The rapid spread of the virus has demonstrated its ability to overwhelm the health systems of developed countries and poses in danger countries with vulnerable health systems. These particular characteristics underline the adoption of social-distancing measures including quarantines, lockdowns and limitations in international travel implemented by most governments.

Until now, the most widely used diagnostic tool is real-time reverse transcriptase polymerase chain reaction (RT-PCR) from nasal and pharyngeal swabs (3). Although, RT-PCR is highly specific, has a suboptimal sensitivity (4), and false negatives have been reported (5). Without proven therapeutic drugs or vaccines, the identification and isolation of infected people through contact tracing or passive detection is of vital importance for controlling disease spread (4). In this context, an adequate characterization of suspected cases becomes of critical importance.

We analyzed the symptoms corresponding to a large database obtained from individuals evaluated by RT-PCR in Argentina in order to identify the symptoms associated with COVID-19 and develop a model to identify possible individuals infected with SARS-CoV-2.

## Materials and Methods

### Data source

The data file was provided by the National Ministry of Health of Argentina (Ministerio de Salud de la Nación Argentina. Dirección Nacional de Epidemiología y Análisis de la Situación de Salud. Área de Vigilancia). The analysis included 67318 people evaluated through RT-PCR for SARS-CoV-2 throughout the country from April 1 through May 24, 2020 and included in the registry of the National Ministry of Health. Several variables including age, sex, RT-PCR result (positive or negative), and symptoms at presentation were recorded in a structured questionnaire for each individual. Symptoms (presence or absence) include anosmia, dysgeusia, arthralgia, headache, confusion, seizures, diarrhea, dyspnea, abdominal pain, chest pain, low grade fever (37.5-37.9° Celsius), high grade fever (≥38°C), respiratory failure, conjunctival injection, irritability, malaise, myalgia, food refusal, tachypnea, use of accessory muscles for breathing (UAMB), cough and vomiting. Before May 18 the form included ‘odynophagia’ and ‘sore throat’. After May 18 only odynophagia remained in the form. Therefore, all odynophagia analyzes were performed considering the data from May 18 to May 24 (10225 individuals - 15 % of the entire dataset), the period of time where only odynophagia was available in the questionnaire. We did not impose any further exclusion criteria to limit selection bias. This study is reported as per the Strengthening the Reporting of Observational Studies in Epidemiology (STROBE) guidelines (S1 STROBE Checklist, Supporting material).

### Diagnosis by RT-PCR

The diagnosis by RT-PCR was used as a reference standard for the design of the regression model. All kits for use in Argentina were approved by the *Administración Nacional de Medicamentos, Alimentos y Tecnología Médica*. A fluid sample was taken with a swab from each nostril and pharynx. From these samples, an RNA extraction was performed for the subsequent RT-PCR.

### Data analysis

Symptoms of COVID-19 have been reported to differ in different age groups (6), therefore two age categories were used for the analysis. A group with ages from 0 to 55 years (<56 group, n = 48748) and another group with ages 56 to 103 years-old (≥56 group, n = 18570). The association of symptoms (presence versus absence) and sex (male versus female) with a RT-PCR positive result for SARS-CoV-2 was studied using multivariate logistic regression analysis considering pairwise interactions, in both age groups. The effect of the different covariates was quantified by the odds ratio values and the corresponding 95% confidence intervals obtained by logistic regression.

### Predictive models

We developed and tested different models aimed to improve the identification of suspected cases; with the premise that such models should have the fewest significant variables and symptoms that could be collected without the need for a physical exam and laboratory testing (e.g. respiratory failure). Odynophagia was excluded from the model design because this symptom alone (that is, without the option ‘sore throat’) was recorded only for 10225 individuals, and the exclusion of this symptom did not produce a loss of the predictive capacity of the models.

To develop and test the models, the database (n=67318) was randomly divided into two data sets (Training and Testing datasets) with the same percentage of RT-PCR positive cases in each one. The Training set (n=53854) was used for the design and internal validation of the models, while the Testing set (n=13464) was used for external validation and evaluation of the predictive capacity of the model. Models were compared using the Akaike Information Criterion, the area under the receiver-operator characteristic curve (AUC) and the predictive value.

The best model was chosen through stepwise regression with bidirectional elimination based on AIC. In addition, we performed a 100-fold cross-validation method in which the training base is partitioned in 100 randomly selected subsets and the model is fitted for each one. All statistical analyzes were performed with R (7), and GraphPad Prism version 5.00 for Windows (La Jolla California USA, www.graphpad.com).

## Results

### Data description

A total of 67318 individuals were included in this study; among them, 12% had a positive RT-PCR for SARS-CoV-2 with a median of 37 years-old (IQR: 26-51), while the group with a negative RT-PCR had a median of 40 years-old (IQR: 25-59). Infection rates varied in different age groups, showing the highest rates in the 15 to 60 years-old group (Figure 1).

**Figure 1:**
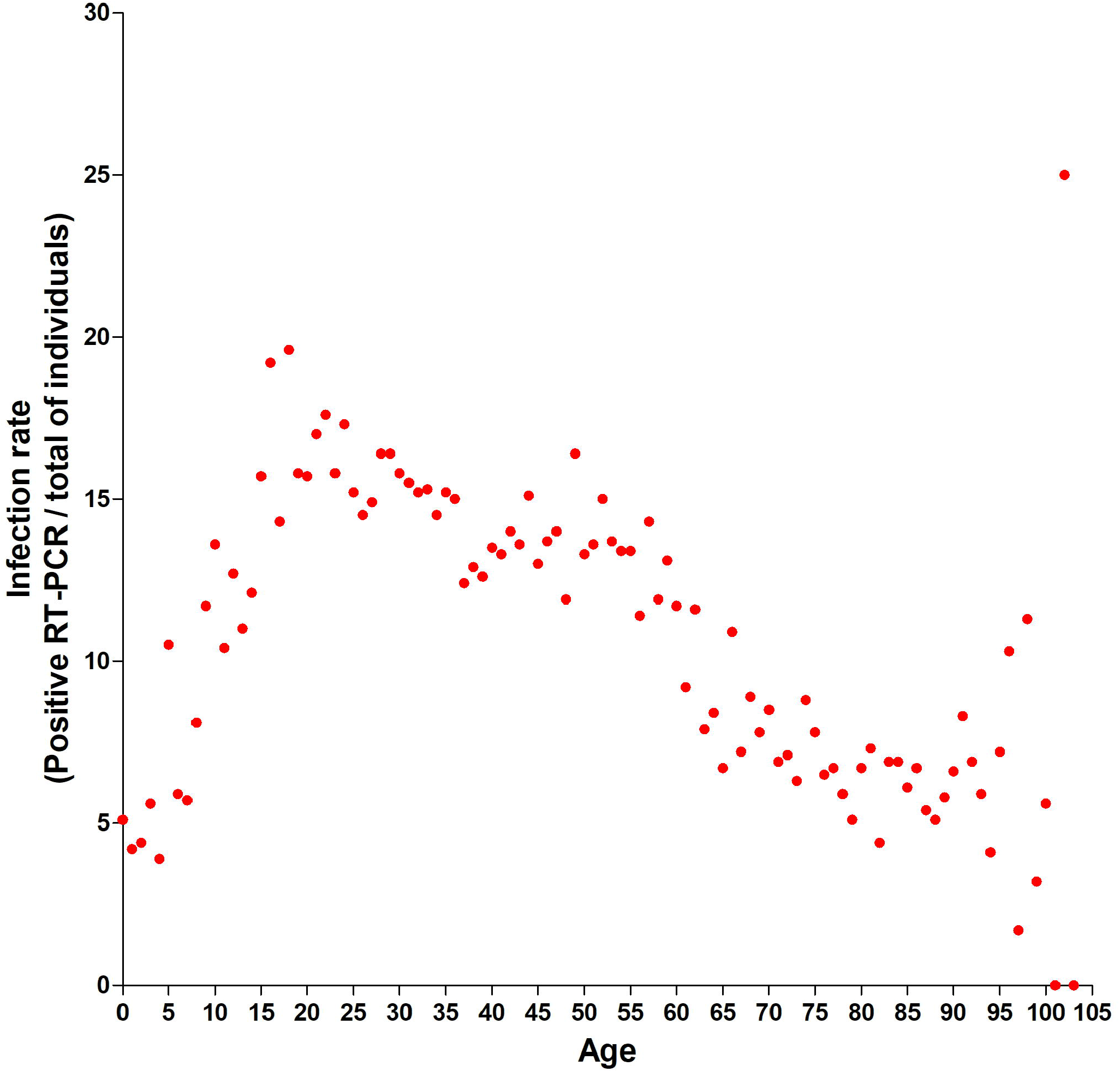
SARS-CoV-2 infection in relation to age.

Significant differences were identified in the positivity rate of RT-PCR according to the interval between the onset of symptoms and the time of sampling. It was observed that for the day ranges of 0-3, 4-15 and 16-30 the proportion of positives was 10.8, 13.7 and 8.6% respectively. Between the three intervals of days a significant difference was observed (p <0.01).

The <56 group (n = 48748) presented a median age of 32 (IQR: 20-42), where 48% were male and 52% female while the median age for the ≥56 group (n=18570) was 72 (IQR: 63-81, 52% male and 48% female).

The most frequent symptoms for each age group were independent of the RT-PCR result (Figure 2). For the <56 group the most frequently observed symptoms were high-grade fever, cough, odynophagia, malaise and headache. For the ≥56 group the most frequent symptoms were high-grade fever, cough, tachypnea, odynophagia and malaise. However, the symptoms with the highest proportion of a confirmatory RT-PCR result were anosmia, dysgeusia, low grade fever, odynophagia, headache, and cough for the <56 group (Figure 3a); and anosmia, dysgeusia, odynophagia and low grade fever for the ≥56 group (Figure 3b).

**Figure 2:**
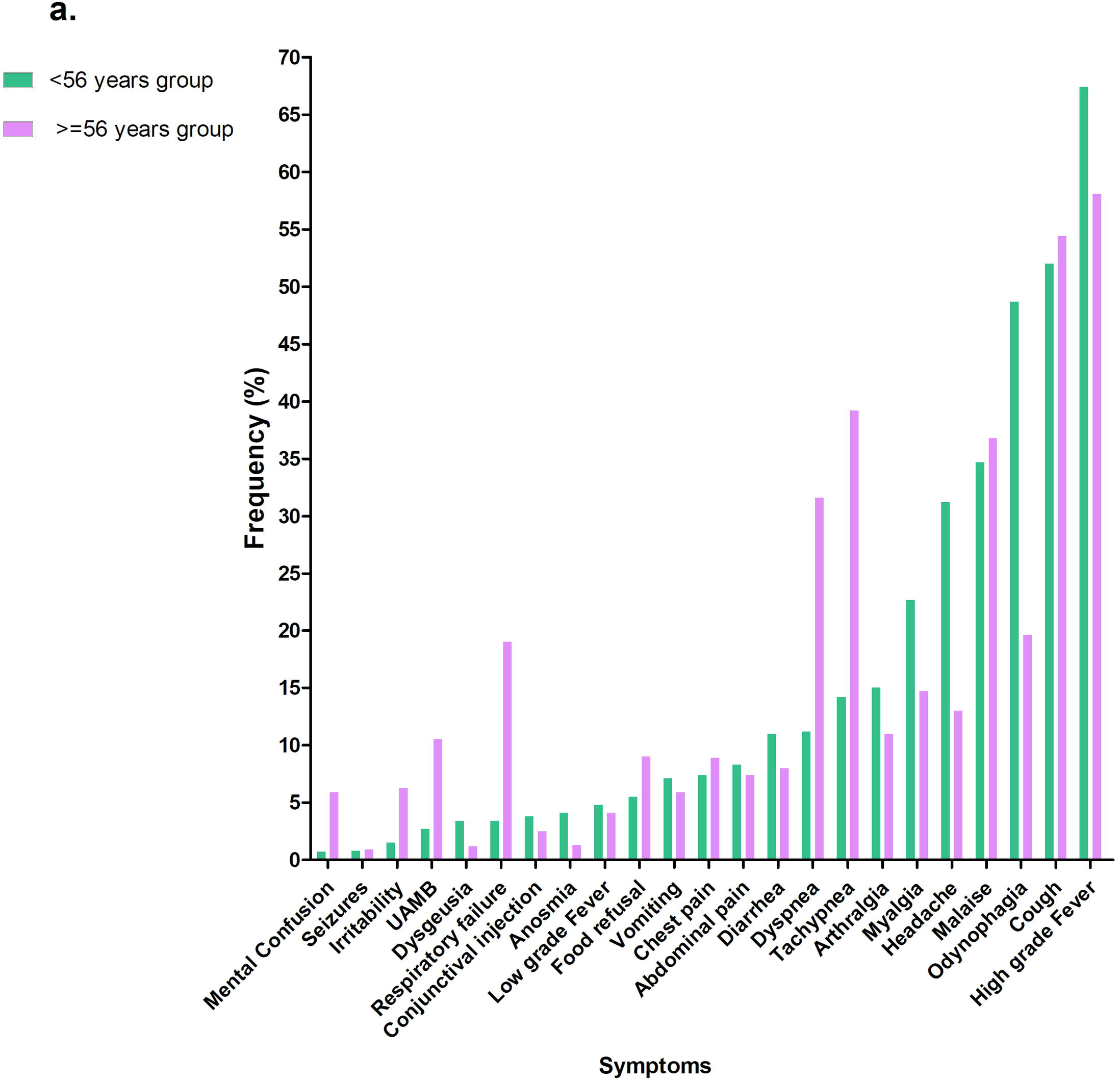
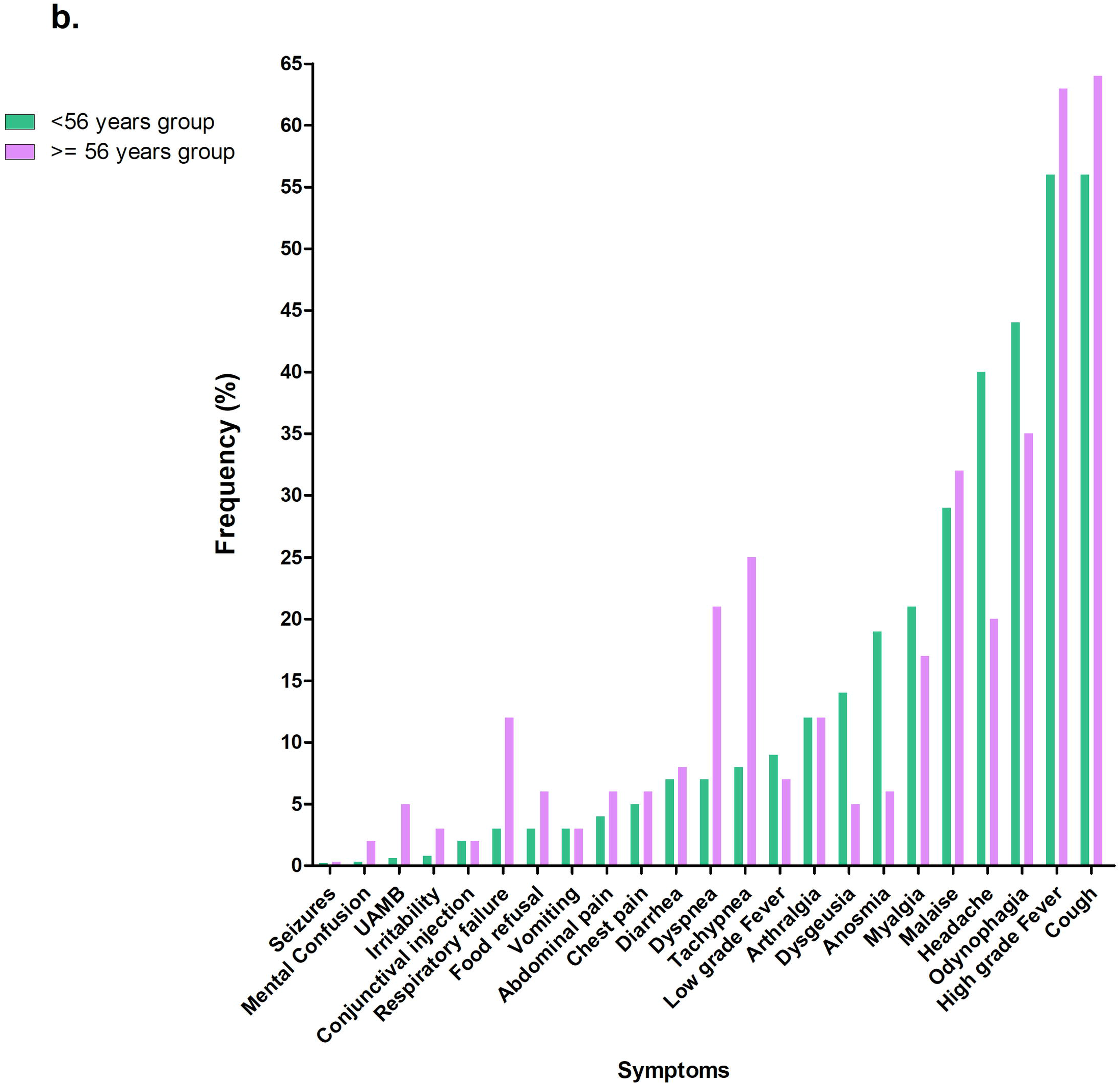
Frequency of symptoms in the <56 group, and ≥56 group. **a**. whole study population with positive and negative RT-PCR. **b**. Only individuals with positive RT-PCR. For odynophagia, the analyzed database is restricted to 10255 individuals.

**Figure 3:**
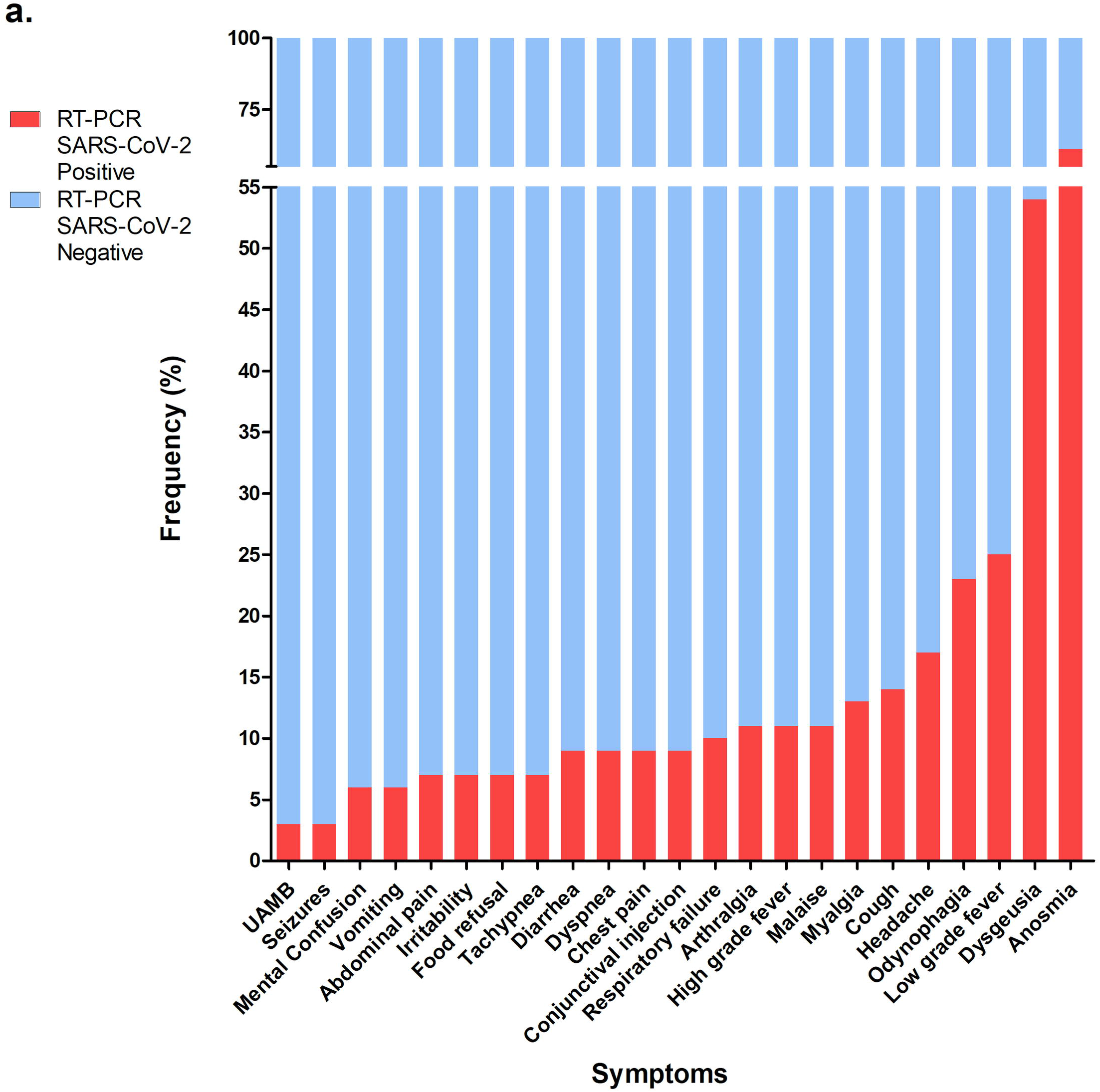
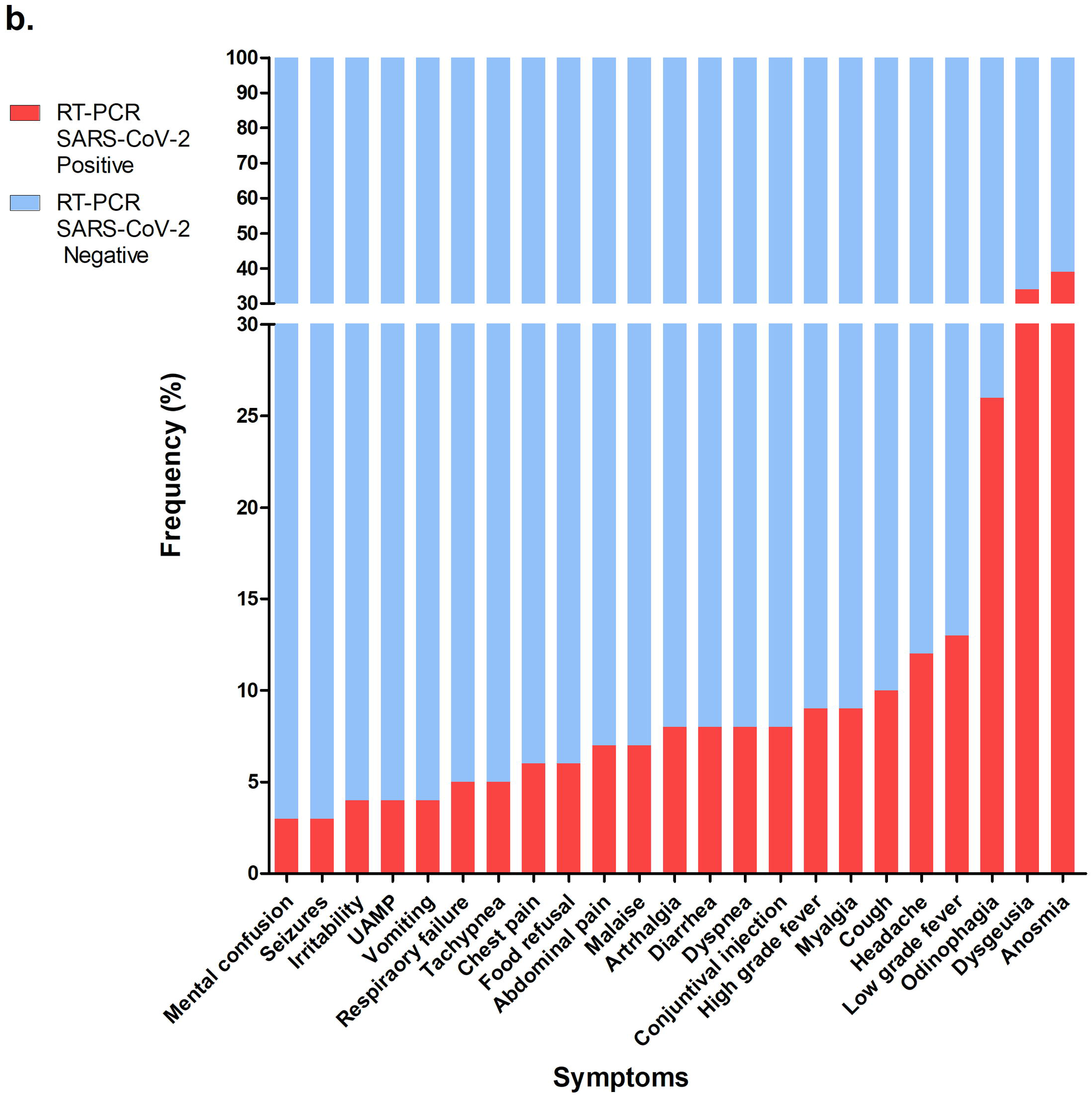
Proportion (%) of individuals with positive and negative RT-PCR for SARS-CoV-2 for each of the symptoms used in the database. **a**. <56 group (n: 48748). **b**. ≥56 group (n: 18570). For odynophagia, the analyzed database is restricted to 10255 individuals.

### Clinical features associated with COVID-19

The association between sex, symptoms and infection with SARS-CoV-2 was studied using logistic regression analysis considering pairwise interactions (see supporting material for details). The odd ratios obtained for each of the symptoms evaluated are displayed in Figure 4 for both age-groups. Sex was significant only for the <56 group with males appearing with higher odds than females (OR =1.23 (IC 95%: 1.07,1.40)).

**Figure 4:**
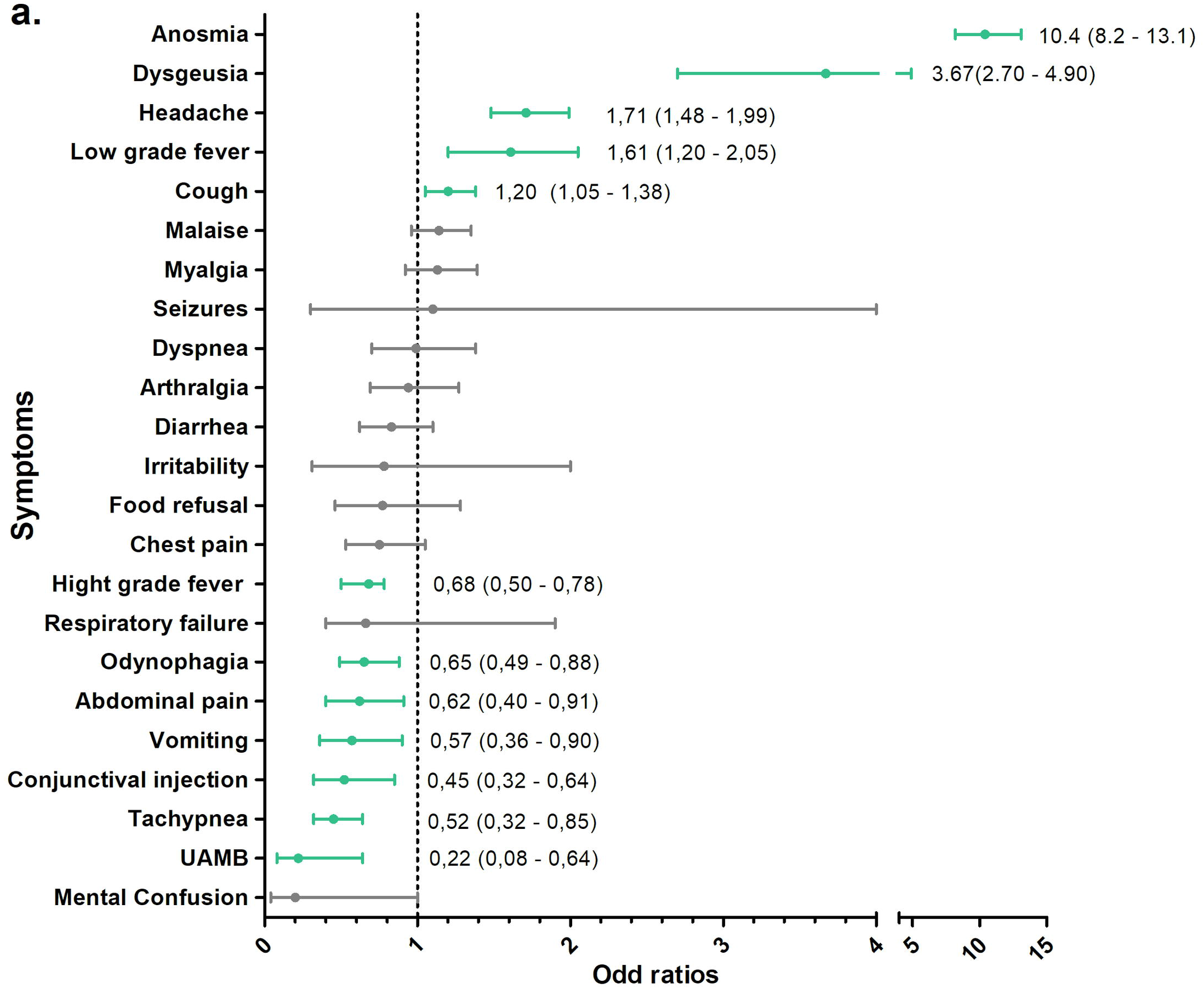
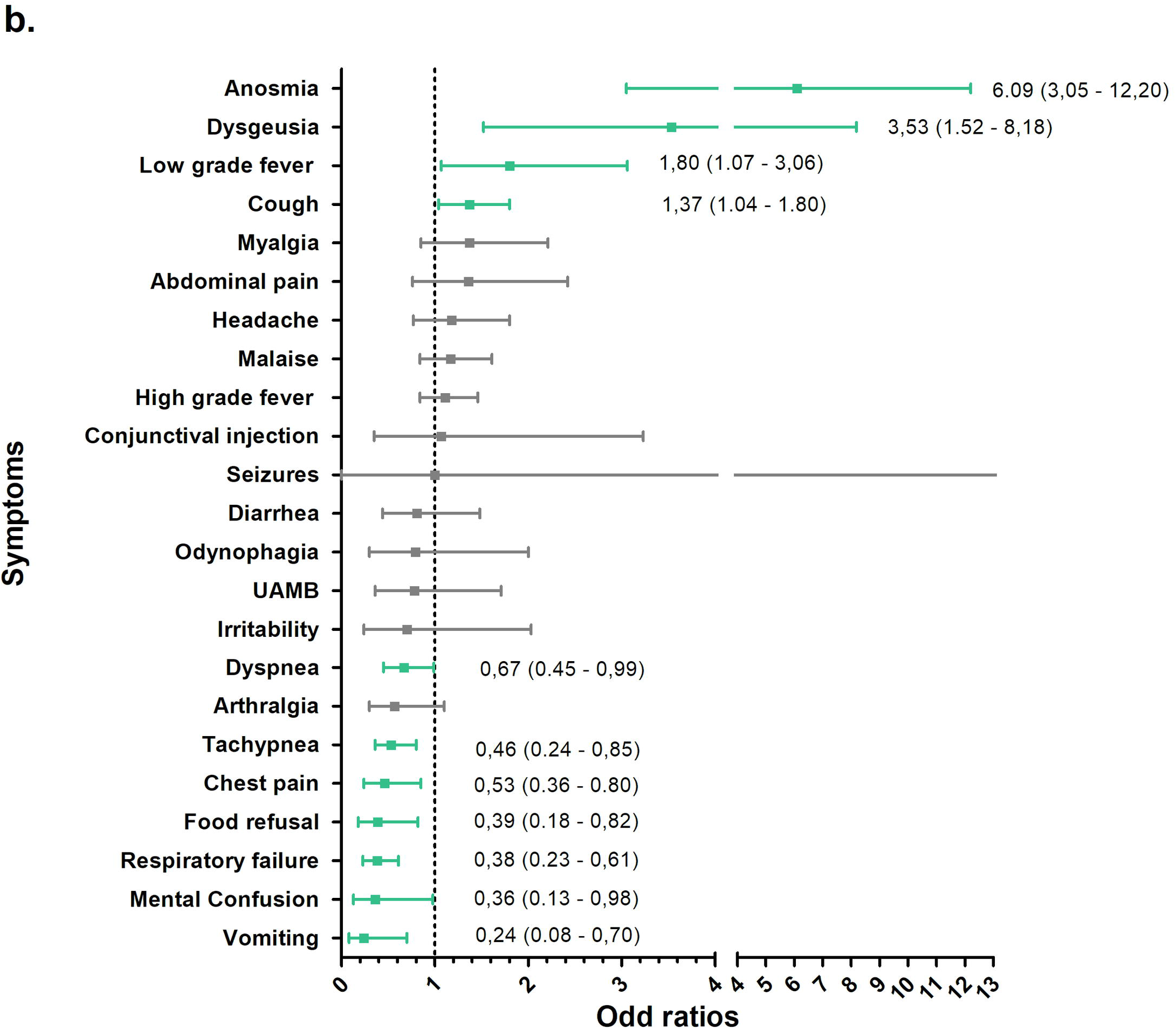
Odd ratios (OR) of SARS-CoV-2 infection corresponding to the presence of symptoms for (a) <56 group and (b) ≥56 group. In green statistically significant ORs (p <0.05) and in gray non-significant ORs (p> 0.05). For odynophagia, the analyzed database is restricted to 10255 individuals.

In the <56 group the symptoms with a positive association with COVID-19 were anosmia, dysgeusia, headache, low grade fever and cough; while UAMB, tachypnea, conjunctival injection, vomiting, abdominal pain, odynophagia and high grade fever have a negative association. The rest of the symptoms did not show statistical significant association with COVID-19, but some of them have a significant interaction with other symptoms (S2 Table, Supporting material). Significantly higher ORs were found with the combination of anosmia and dysgeusia (OR=19.84), anosmia and conjunctival injection (OR=17.40), anosmia and cough (OR=9.73) and anosmia and malaise (OR=6.14).

For the ≥56 group, symptoms at presentation with positive association with COVID-19 were anosmia, disgeusia, low grade fever and cough, while vomiting, respiratory failure, food refusal, chest pain, tachypnea and dyspnea presented negative association. Other symptoms did not show statistically significant association with COVID-19, but as in the previous case, some have significant interactions (S3 Table, Supporting material). Significantly higher ORs were found with the combination of low-grade fever and irritability (OR=14.32), and anosmia and dysgeusia (OR=6.44).

### Logistic regression predictive model

We considered several different logistic models for the probability of SARS-CoV-2 infection:

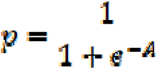

where A depends of the set of symptoms considered and other variables like sex, age or days since symptoms onset. All tested models presented similar characteristics regarding area under the curve, AIC value and predictive value. The model with lower AIC value (35029) considers 16 symptoms, sex and three quantitative variables: age, age squared and the number of symptoms (NS) as described in Table 1. Cross validation did not show significant differences between the model calculated with the training set and 100 randomly chosen partitions. The variable age square captures the behavior of the proportion of positive cases with age (Figure 1). An alternative model without the variable age square is described in the supporting material S4 Appendix. It can also be observed that the number of symptoms has a negative contribution in the equation resulting in decreases in the likelihood of COVID-19 infection as the number of symptoms increases.

**Table 1:** Characteristics of the logistic regression model. p: probability of being infected with SARS-CoV-2, AUC= Area under the receiver-operator characteristic curve, PPV: predicted positive value, NPV: Negative predicted value.

| Equation<br>$P=1/(1+e^{-A})$<br>AUC=0.7167 | Predictive capacity for a cut line of $p=0.08839$ |
| --- | --- |
| $A = -2.137 + 0.03422 \cdot \text{age} - 0.001131 \cdot \text{age}^2 - 0.7953 \cdot \text{AS} - 0.1805 \cdot \text{sex} + 2.236 \cdot \text{anosmia} + 0.2680 \cdot \text{arthralgia} - 0.5870 \cdot \text{seizure} + 0.1484 \cdot \text{diarrhea} + 1.272 \cdot \text{dysgeusia} + 0.1812 \cdot \text{dyspnea} + 0.01535 \cdot \text{chest paint} + 0.9989 \cdot \text{low fever} + 0.2446 \cdot \text{high fever} + 0.08885 \cdot \text{conjunctival injection} + 0.2330 \cdot \text{discomfort} + 0.4379 \cdot \text{myalgia} + 0.2694 \cdot \text{food refusal} - 0.3359 \cdot \text{tachypnea} + 1.066 \cdot \text{cough} + 1.640 \cdot \text{headache} + 7.964 \cdot 10^{-6} \cdot \text{age} \cdot \text{age}^2 + 0.01485 \cdot \text{age} \cdot \text{AS} + 0.01807 \cdot \text{age} \cdot \text{sex} - 0.03391 \cdot \text{age} \cdot \text{headache} - 0.01412 \cdot \text{age} \cdot \text{cough} - 1.178 \cdot 10^{-4} \cdot \text{age}^2 \cdot \text{AS} - 1.981 \cdot 10^{-4} \cdot \text{age}^2 \cdot \text{sex} + 2.596 \cdot 10^{-4} \cdot \text{age}^2 \cdot \text{headache} + 1.446 \cdot 10^{-4} \cdot \text{age}^2 \cdot \text{cough} + 0.7360 \cdot \text{anosmia} \cdot \text{conjunctival injection} + 0.6033 \cdot \text{diarrhea} \cdot \text{chest paint} + 0.7430 \cdot \text{dysgeusia} \cdot \text{tachypnea} + 0.6774 \cdot \text{dyspnea} \cdot \text{conjunctival injection} + 0.4194 \cdot \text{high fever} \cdot \text{tachypnea}$ | Sensitivity=0.80<br>(0.78,0.82) |
|  | Specificity=0.46<br>(0.45,0.47) |
|  | PPV=0.17 (0.16,0.17) |
|  | NPV=0.95 (0.94,0.95) |

## Discussion

The analysis of this large cohort allows an age-based characterization of the symptoms associated with COVID-19 at presentation, in the search for an improved clinical based decision strategy for the use of diagnostic tests and case management. To interrupt the spread of SARS-CoV-2, early detection, isolation, and the implementation of a robust system to trace contacts is required (WHO, 2020). Among the non-severe symptomatic cases, non-specific symptoms shared with other acute upper-respiratory infections hamper proper case identification and implementation of rapid isolation measures. In contrast to most acute upper-respiratory infections (including many respiratory coronaviruses), COVID-19 poses a special challenge in requiring specific identification for clinical and public health management. To facilitate the early detection of patients with COVID-19, we analyzed the symptoms of 67318 individuals studied with RT-PCR, of which 12% had a positive test result.

The most frequently reported symptoms among case series of COVID-19 include fever, cough, dyspnea and shortness of breath (8,9). Other symptoms include sore throat, diarrhea, nausea, vomiting, myalgia, headache and confusion (8). We found that in the present database the most frequent symptoms were cough, high-grade fever, and odynophagia. However, more frequent symptoms not necessarily are indicators of COVID-19. Our analysis revealed that the symptoms positively associated with COVID-19 are anosmia, dysgeusia, low grade fever (37.5 to 38 °C), cough and headache (the later only in the <56 group). Another group of symptoms show a negative association with a positive RT-PCR for SARS-CoV-2, therefore suggesting alternative diagnoses (Figure 3). Our analysis also revealed that some symptoms interact with each other, modifying their ORs. In relation to this, anosmia and dysgeusia were observed to have a significant and positive interaction increasing the already high and positive OR of each of them. This is in accordance with other authors stating that anosmia and dysgeusia have a positive predictive value of 77% each of them and 83% if they are together (10). Although anosmia can appear in other AURI, it has been suggested that the lack of accompanying rhinitis or nasal swelling is more typically associated with COVID-19 (11); we were not able to test this hypothesis because the questionnaire does not contain rhinitis as a variable. Another combined effect to be highlighted was anosmia and conjunctival injection in the<56 age group and low-grade fever and irritability in the elder group. These combinations showed ORs >14. Low grade fever had a positive association with COVID-19 in both age groups, while high grade fever (≥38°C) showed a negative association in the <56 group and was not significant in the ≥56 group.

In this study we also developed a model based on symptoms at presentation, sex and age. In our database, patient’s age was found as a significant variable for establishing correlations between symptoms and COVID-19. Although the age-dependent variability in the proportion of RT-PCR positive results in our study population might reflects differences in exposure due to school closure and an active protection of the elder, the model achieves comparable predictive values in all age groups. In view of the burden of morbidity and mortality in high risk groups, models to predict the likelihood of infection should be interpreted with caution and not be used to limit the corresponding testing of groups at increased risk of severe disease.

This study was conducted with patients diagnosed between April and May 2020, prior to the influenza season in Argentina; this should also place our results in the adequate context of the alternative diagnoses that could modify the specificity of the symptoms based on the type and prevalence of these agents; in the current case, with most cases occurring in Buenos Aires and its metropolitan area, this same area was affected by its largest recorded dengue outbreak and a significant reduction of influenza-like illnesses (12).

This study has limitations; the diagnostic test of choice, RT-PCR shows a sensitivity of 56–83% (13); therefore, the sensitivity and specificity measures of the model may lose precision. While suffering from the lack of a highly sensitive diagnostic standard, identifying clinical characteristics that raise the pretest probability of the infection could help interpretations of the RT-PCR result when combined with predictive models, epidemiological factors and complementary diagnostic methods like chest imaging (14). In our analysis, a significantly higher probability of a positive RT-PCR was identified if the test was performed when the interval between symptoms initiation and sampling was between 4 and 15 days. Another limitation derives from the circumstance of the data collection, which has not been design as a prospective research tool but rather as a surveillance system with no monitoring system and therefore subject to errors in upload.

In summary, this symptoms-based analysis of a cohort tested for COVID-19 identified a group of symptoms (anosmia/dysgeusia, low-grade fever, cough and headache) with significant association with a positive RT-PCR. These findings show that a regression model based on multiple factors (age, sex, interaction between symptoms) could be used as a complementary method for the rapid identification of possible COVID-19 cases and the necessary precautionary measures.

## Data Availability

The data files is available from the website of National Ministry of Health of Argentina

http://datos.salud.gob.ar/dataset/covid-19-determinaciones-registradas-en-la-republica-argentina

## Supporting information captions

- S1 STROBE Checklist.
- S2 Table. Logistic regression for the group with ages from 0 to 55 years (<56 group, n=48748).
- S3 Table. Logistic regression for the group with ages from 56 to 103 years-old (≥56 group, n = 18570).
- S4 Appendix. Alternative model with the variables sex, age, 20 symptoms and pairwise interactions.

## References

1. World Health Organization (WHO). Novel Coronavirus (2019-nCoV) Situation Report - 12 1 February 2020. WHO Bull. 2020;(JANUARY):1–7.

2. Hopkins C for SS and E (CSSE) J. COVID-19 Map - Johns Hopkins Coronavirus Resource Center. [cited 2020 September 5]. Available from: https://coronavirus.jhu.edu/map.html

3. Pascarella G, Strumia A, Piliego C, Bruno F, Del Buono R, Costa F, et al. COVID-19 diagnosis and management: a comprehensive review. Vol. 2019, Journal of Internal Medicine. 2020. 0–2 p.

4. Ai T, Yang Z, Hou H, Zhan C, Chen C, Lv W, et al. Correlation of Chest CT and RT-PCR Testing in Coronavirus Disease 2019 (COVID-19) in China: A Report of 1014 Cases. 2020;78(May):1–15.

5. Zhang JJ, Cao YY, Dong X, Wang BC, Liao MY, Lin J, et al. Distinct characteristics of COVID-19 patients with initial rRT-PCR positive and negative results for SARS-CoV-2. Allergy. 2020.

6. Yang R, Gui X, Xiong Y. Comparison of Clinical Characteristics of Patients with Asymptomatic vs Symptomatic Coronavirus Disease 2019 in Wuhan, China. JAMA Netw open. 2020;3(5):e2010182.

7. R Core Team. R: A language and environment for statistical computing. R Foundation for Statistical Computing, Vienna, Austria. 2013. Available from: http://www.r-project.org/

8. Wu YC, Chen CS, Chan YJ. The outbreak of COVID-19: An overview. J Chinese Med Assoc. 2020;83(3):217–20.

9. Yang J, Zheng Y, Gou X, Pu K, Chen Z, Guo Q, et al. Prevalence of comorbidities and its effects in coronavirus disease 2019 patients: A systematic review and meta-analysis. Int J Infect Dis. 2020;94:91–5.

10. Zayet S, Klopfenstein T, Mercier J, Kadiane-Oussou NJ, Lan Cheong Wah L, Royer P-Y, et al. Contribution of anosmia and dysgeusia for diagnostic of COVID-19 in outpatients. Infection. 2020;(0123456789):0–4.

11. Zubair AS, McAlpine LS, Gardin T, Farhadian S, Kuruvilla DE, Spudich S. Neuropathogenesis and Neurologic Manifestations of the Coronaviruses in the Age of Coronavirus Disease 2019: A Review. JAMA Neurology. American Medical Association; 2020.

12. Boletín integral de vigilancia. Edición semanal. 499 Boletín Integrado de Vigilancia. 2020 [cited 2020 Jul 7]. Available from: https://www.argentina.gob.ar/sites/default/files/biv_499_se23.pdf

13. Kokkinakis I, Selby K, Favrat B, Genton B, Cornuz J. [Covid-19 diagnosis□: clinical recommendations and performance of nasopharyngeal swab-PCR]. Rev Med Suisse. 2020;16(689):699—701.

14. Woloshin S, Patel N, Kesselheim AS. False Negative Tests for SARS-CoV-2 Infection — Challenges and Implications. N Engl J Med. 2020 Jun 5.

