## Supplementary material for "Understanding the value of clinical symptoms of COVID-19. A logistic regression model": S1 STROBE Checklist.pdf

STROBE Statement—Checklist of items that should be included in reports of cross-sectional studies

| | Item No. | Recommendation | Page No. | Relevant text from manuscript |
|---|---|---|---|---|
| **Title and abstract** | 1 | (*a*) Indicate the study's design with a commonly used term in the title or the abstract | Abstract, Page 3 | "*cross sectional study*" |
| | | (*b*) Provide in the abstract an informative and balanced summary of what was done and what was found | Abstract, Pages 3 and 4 | |
| **Introduction** | | | | |
| Background/rationale | 2 | Explain the scientific background and rationale for the investigation being reported | Introduction, Pages 5 | |
| Objectives | 3 | State specific objectives, including any prespecified hypotheses | Introduction, Page 5 | "*We analysed the symptoms corresponding to a large database obtained from individuals evaluated by RT-PCR in Argentina in order to identify the symptoms associated with COVID-19 and develop a model to identify possible individuals infected with SARS-CoV-2..*" |
| **Methods** | | | | |
| Study design | 4 | Present key elements of study design early in the paper | Methods, Page 6 | |
| Setting | 5 | Describe the setting, locations, and relevant dates, including periods of recruitment, exposure, follow-up, and data collection | Methods, Page 6 | |
| Participants | 6 | Give the eligibility criteria, and the sources and methods of selection of participants | Methods, Page 6 | *"The analysis included 67318 people evaluated through RT-PCR for SARS-CoV-2 throughout the country from April 1 through May 24, 2020 and included in the registry of the National Ministry of Health.* |

| | | | | |
|---|---|---|---|---|
| | | | | *Several variables including age, sex, RT-PCR result (positive or negative), and symptoms at presentation were recorded in a structured questionnaire for each individual."* |
| Variables | 7 | Clearly define all outcomes, exposures, predictors, potential confounders, and effect modifiers. Give diagnostic criteria, if applicable | Methods, Page 7 | "*The association of symptoms (presence versus absence) and sex (male versus female) with a RT-PCR positive result for SARS-CoV-2 was studied using multivariate logistic regression analysis considering pairwise interactions*" |
| Data sources/ measurement | 8* | For each variable of interest, give sources of data and details of methods of assessment (measurement). Describe comparability of assessment methods if there is more than one group | Methods, Pages 6 and 7 | *"Several variables including age, sex, RT-PCR result (positive or negative), and symptoms at presentation were recorded in a structured questionnaire for each individual. Symptoms (presence or absence) include…"* *"two age categories were used for the analysis. A group with ages from 0 to 55 years (<56 group, n = 48748) and another group with ages 56 to 103 years-old (≥56 group, n = 18570)."* |

| | | | | |
|---|---|---|---|---|
| Bias | 9 | Describe any efforts to address potential sources of bias | Methods, Page 6 | "*We did not impose any further exclusion criteria to limit selection bias.*" |
| Study size | 10 | Explain how the study size was arrived at | Methods, Page 6 | |

Continued on next page

| | | | | |
|---|---|---|---|---|
| Quantitative variables | 11 | Explain how quantitative variables were handled in the analyses. If applicable, describe which groupings were chosen and why | Methods, Page 7 | |
| Statistical methods | 12 | (*a*) Describe all statistical methods, including those used to control for confounding | Methods, Pages 7 and 8 | |
| | | (*b*) Describe any methods used to examine subgroups and interactions | Methods, Page 7 | |
| | | (*c*) Explain how missing data were addressed | Methods, Page 6 | “ *Before May 18 the form included ‘odynophagia’ and ‘sore throat’. After May 18 only odynophagia remained in the form. Therefore, all odynophagia analyzes were performed considering the data from May 18 to May 24 (10225 individuals - 15 % of the entire dataset), the period of time where only odynophagia was available in the questionnaire*.” |
| | | (*d*) If applicable, describe analytical methods taking account of sampling strategy | N/a | |
| | | (*e*) Describe any sensitivity analyses | Methods, Pages 7 and 8 | |
| **Results** | | | | |
| Participants | 13* | (a) Report numbers of individuals at each stage of study—eg numbers potentially eligible, examined for eligibility, confirmed eligible, included in the study, completing follow-up, and analysed | Results, Page 9 | “*A total of 67318 individuals were included in this study; among them, 12% had a positive RT-PCR for SARS-CoV-2*” |
| | | (b) Give reasons for non-participation at each stage | N/a | |
| | | (c) Consider use of a flow diagram | N/a | |
| Descriptive data | 14* | (a) Give characteristics of study participants (eg demographic, clinical, social) and information on exposures and potential confounders | Results, Page 9 | |

| | | | |
|---|---|---|---|
| | | (b) Indicate number of participants with missing data for each variable of interest | N/a |
| Outcome data | 15* | *Cross-sectional study*—Report numbers of outcome events or summary measures | Results, Page 9, Figure 2 and Figure 3 |
| Main results | 16 | (*a*) Give unadjusted estimates and, if applicable, confounder-adjusted estimates and their precision (eg, 95% confidence interval). Make clear which confounders were adjusted for and why they were included | Figure 4, S1 Table and S2 Table |
| | | (*b*) Report category boundaries when continuous variables were categorized | N/a |
| | | (*c*) If relevant, consider translating estimates of relative risk into absolute risk for a meaningful time period | N/a |

Continued on next page

| | | | | |
|---|---|---|---|---|
| Other analyses | 17 | Report other analyses done—eg analyses of subgroups and interactions, and sensitivity analyses | Table 1 and S3 Appendix | |
| **Discussion** | | | | |
| Key results | 18 | Summarise key results with reference to study objectives | Discussion, Page 14 | |
| Limitations | 19 | Discuss limitations of the study, taking into account sources of potential bias or imprecision. Discuss both direction and magnitude of any potential bias | Discussion, Page 16 | |
| Interpretation | 20 | Give a cautious overall interpretation of results considering objectives, limitations, multiplicity of analyses, results from similar studies, and other relevant evidence | Discussion Pages 15 and 16 | "*In view of the burden of morbidity and mortality in high risk groups, models to predict the likelihood of infection should be interpreted with caution and not be used to limit the corresponding testing of groups at increased risk of severe disease*" |
| Generalisability | 21 | Discuss the generalisability (external validity) of the study results | Discussion, Pages 15 and 16 | |
| **Other information** | | | | |
| Funding | 22 | Give the source of funding and the role of the funders for the present study and, if applicable, for the original study on which the present article is based | N/a | |

*Give information separately for cases and controls in case-control studies and, if applicable, for exposed and unexposed groups in cohort and cross-sectional studies.

**Note:** An Explanation and Elaboration article discusses each checklist item and gives methodological background and published examples of transparent reporting. The STROBE checklist is best used in conjunction with this article (freely available on the Web sites of PLoS Medicine at http://www.plosmedicine.org/, Annals of Internal Medicine at http://www.annals.org/, and Epidemiology at http://www.epidem.com/). Information on the STROBE Initiative is available at www.strobe-statement.org.
