## Supplementary material for "Understanding the value of clinical symptoms of COVID-19. A logistic regression model": S3 Table.pdf

**S3 Table. Logistic regression for the group with ages from 56 to 103 years-old (≥56 group, n = 18570).** Odynophagia was evaluated only in 2370 individuals. High_fever= High grade fever, Low_fever= Low grade fever, M=male, 1= symptom present. Variables with statistical significance (p <0.05) are highlighted in yellow.

| Variable | Adjusted Odd Ratio | Lower 95% CI | Upper 95% CI | p |
|---|---|---|---|---|
| Sex(M) | 8.99e-01 | 6.81e-01 | 1.19e+00 | 4.50e-01 |
| Anosmia(1) | 6.09e+00 | 3.05e+00 | 1.22e+01 | 3.23e-07 |
| Arthralgia(1) | 5.72e-01 | 3.00e-01 | 1.09e+00 | 8.96e-02 |
| Headache(1) | 1.18e+00 | 7.74e-01 | 1.80e+00 | 4.40e-01 |
| Mental_confusion(1) | 3.55e-01 | 1.29e-01 | 9.78e-01 | 4.53e-02 |
| Seizures(1) | 6.85e-20 | 0.00e+00 | Inf | 9.41e-01 |
| Diarrhea(1) | 8.11e-01 | 4.43e-01 | 1.49e+00 | 4.97e-01 |
| Dysgeusia(1) | 3.53e+00 | 1.53e+00 | 8.18e+00 | 3.22e-03 |
| Dyspnea(1) | 6.65e-01 | 4.48e-01 | 9.87e-01 | 4.29e-02 |
| Abdominal_pain(1) | 1.36e+00 | 7.60e-01 | 2.42e+00 | 3.02e-01 |
| Chest_pain(1) | 4.57e-01 | 2.45e-01 | 8.55e-01 | 1.42e-02 |
| Low_fever(1) | 1.81e+00 | 1.07e+00 | 3.06e+00 | 2.69e-02 |
| High_fever(1) | 1.11e+00 | 8.44e-01 | 1.47e+00 | 4.47e-01 |
| Respiratory_failure(1) | 3.78e-01 | 2.33e-01 | 6.12e-01 | 7.77e-05 |
| Conjunctival_injection(1) | 1.07e+00 | 3.54e-01 | 3.24e+00 | 9.03e-01 |
| Irritability(1) | 6.99e-01 | 2.41e-01 | 2.03e+00 | 5.11e-01 |
| Malaise(1) | 1.17e+00 | 8.42e-01 | 1.61e+00 | 3.55e-01 |
| Myalgia(1) | 1.37e+00 | 8.49e-01 | 2.22e+00 | 1.96e-01 |
| Food_refusal(1) | 3.88e-01 | 1.85e-01 | 8.16e-01 | 1.25e-02 |
| Tachypnea(1) | 5.38e-01 | 3.64e-01 | 7.95e-01 | 1.87e-03 |
| UAMB(1) | 7.84e-01 | 3.59e-01 | 1.71e+00 | 5.41e-01 |
| Cough(1) | 1.37e+00 | 1.04e+00 | 1.81e+00 | 2.54e-02 |
| Vomiting(1) | 2.42e-01 | 8.35e-02 | 7.02e-01 | 8.98e-03 |
| Odynophagia(1) | 7.93e-01 | 3.04e-01 | 2.07e+00 | 6.35e-01 |
| Sex(M):Anosmia(1) | 6.26e-01 | 3.07e-01 | 1.28e+00 | 1.98e-01 |
| Sex(M):Arthralgia(1) | 1.06e+00 | 7.13e-01 | 1.57e+00 | 7.76e-01 |
| Sex(M):Headache(1) | 1.59e+00 | 1.17e+00 | 2.16e+00 | 3.06e-03 |
| Sex(M):Mental_confusion(1) | 5.81e-01 | 2.47e-01 | 1.37e+00 | 2.15e-01 |
| Sex(M):Seizures(1) | 1.44e+05 | 6.70e-149 | 3.10e+158 | 9.47e-01 |
| Sex(M):Diarrhea(1) | 7.42e-01 | 4.79e-01 | 1.15e+00 | 1.79e-01 |
| Sex(M):Dysgeusia(1) | 1.85e+00 | 8.48e-01 | 4.05e+00 | 1.22e-01 |
| Sex(M):Dyspnea(1) | 8.57e-01 | 6.32e-01 | 1.16e+00 | 3.20e-01 |
| Sex(M):Abdominal_pain(1) | 9.63e-01 | 5.91e-01 | 1.57e+00 | 8.79e-01 |
| Sex(M):Chest_pain(1) | 7.95e-01 | 5.00e-01 | 1.26e+00 | 3.33e-01 |
| Sex(M):Low_fever(1) | 1.00e+00 | 6.05e-01 | 1.66e+00 | 9.89e-01 |
| Sex(M):High_fever(1) | 9.20e-01 | 7.18e-01 | 1.18e+00 | 5.07e-01 |
| Sex(M):Respiratory_failure(1) | 1.45e+00 | 1.00e+00 | 2.10e+00 | 4.88e-02 |
| Sex(M):Conjunctival_injection(1) | 1.66e+00 | 7.34e-01 | 3.77e+00 | 2.22e-01 |
| Sex(M):Irritability(1) | 9.94e-01 | 4.54e-01 | 2.18e+00 | 9.89e-01 |
| Sex(M):Malaise(1) | 1.05e+00 | 8.12e-01 | 1.35e+00 | 7.28e-01 |

| | | | | |
|---|---|---|---|---|
| Sex(M):Myalgia(1) | 1.00e+00 | 7.15e-01 | 1.41e+00 | 9.81e-01 |
| Sex(M):Food_refusal(1) | 1.13e+00 | 7.04e-01 | 1.82e+00 | 6.09e-01 |
| Sex(M):Tachypnea(1) | 1.12e+00 | 8.36e-01 | 1.50e+00 | 4.45e-01 |
| Sex(M):UAMB(1) | 6.56e-01 | 3.81e-01 | 1.13e+00 | 1.28e-01 |
| Sex(M):Cough(1) | 1.19e+00 | 9.48e-01 | 1.50e+00 | 1.32e-01 |
| Sex(M):Vomiting(1) | 1.05e+00 | 5.45e-01 | 2.02e+00 | 8.88e-01 |
| Anosmia(1):Arthralgia(1) | 6.19e-01 | 1.67e-01 | 2.29e+00 | 4.73e-01 |
| Anosmia(1):Headache(1) | 9.38e-01 | 4.20e-01 | 2.09e+00 | 8.76e-01 |
| Anosmia(1):Mental_confusion(1) | 2.43e+00 | 0.00e+00 | Inf | 1.00e+00 |
| Anosmia(1):Seizures(1) | 7.53e+13 | 0.00e+00 | Inf | 9.96e-01 |
| Anosmia(1):Diarrhea(1) | 1.07e+00 | 2.28e-01 | 5.06e+00 | 9.28e-01 |
| Anosmia(1):Dysgeusia(1) | 3.05e-01 | 1.35e-01 | 6.86e-01 | 4.09e-03 |
| Anosmia(1):Dyspnea(1) | 3.93e-01 | 8.67e-02 | 1.78e+00 | 2.27e-01 |
| Anosmia(1):Abdominal_pain(1) | 4.97e-01 | 1.04e-01 | 2.39e+00 | 3.83e-01 |
| Anosmia(1):Chest_pain(1) | 2.52e+00 | 3.63e-01 | 1.74e+01 | 3.50e-01 |
| Anosmia(1):Low_fever(1) | 2.45e+00 | 5.12e-01 | 1.17e+01 | 2.62e-01 |
| Anosmia(1):High_fever(1) | 1.78e+00 | 8.32e-01 | 3.83e+00 | 1.37e-01 |
| Anosmia(1):Respiratory_failure(1) | 2.40e+00 | 4.86e-01 | 1.18e+01 | 2.83e-01 |
| Anosmia(1):Conjunctival_injection(1) | 1.74e+00 | 1.48e-01 | 2.04e+01 | 6.61e-01 |
| Anosmia(1):Malaise(1) | 6.01e-01 | 2.47e-01 | 1.46e+00 | 2.61e-01 |
| Anosmia(1):Myalgia(1) | 1.31e+00 | 4.96e-01 | 3.48e+00 | 5.82e-01 |
| Anosmia(1):Food_refusal(1) | 5.50e-01 | 9.13e-02 | 3.31e+00 | 5.13e-01 |
| Anosmia(1):Tachypnea(1) | 8.34e-01 | 2.26e-01 | 3.08e+00 | 7.86e-01 |
| Anosmia(1):UAMB(1) | 7.71e-08 | 0.00e+00 | Inf | 9.95e-01 |
| Anosmia(1):Cough(1) | 1.19e+00 | 5.77e-01 | 2.47e+00 | 6.32e-01 |
| Anosmia(1):Vomiting(1) | 6.91e-08 | 0.00e+00 | Inf | 9.95e-01 |
| Arthralgia(1):Headache(1) | 9.81e-01 | 6.38e-01 | 1.51e+00 | 9.29e-01 |
| Arthralgia(1):Mental_confusion(1) | 1.47e+00 | 2.62e-01 | 8.29e+00 | 6.61e-01 |
| Arthralgia(1):Seizures(1) | 9.78e-01 | 1.41e-03 | 6.77e+02 | 9.95e-01 |
| Arthralgia(1):Diarrhea(1) | 6.50e-01 | 3.51e-01 | 1.20e+00 | 1.69e-01 |
| Arthralgia(1):Dysgeusia(1) | 1.64e+00 | 3.98e-01 | 6.74e+00 | 4.94e-01 |
| Arthralgia(1):Dyspnea(1) | 1.41e+00 | 8.18e-01 | 2.44e+00 | 2.15e-01 |
| Arthralgia(1):Abdominal_pain(1) | 5.59e-01 | 2.44e-01 | 1.28e+00 | 1.68e-01 |
| Arthralgia(1):Chest_pain(1) | 3.09e+00 | 1.62e+00 | 5.91e+00 | 6.30e-04 |
| Arthralgia(1):Low_fever(1) | 1.48e+00 | 5.06e-01 | 4.32e+00 | 4.75e-01 |
| Arthralgia(1):High_fever(1) | 1.55e+00 | 9.64e-01 | 2.50e+00 | 7.03e-02 |
| Arthralgia(1):Respiratory_failure(1) | 5.69e-01 | 2.51e-01 | 1.29e+00 | 1.77e-01 |
| Arthralgia(1):Conjunctival_injection(1) | 6.59e-01 | 2.41e-01 | 1.80e+00 | 4.17e-01 |
| Arthralgia(1):Irritability(1) | 6.59e-01 | 1.47e-01 | 2.94e+00 | 5.85e-01 |
| Arthralgia(1):Malaise(1) | 1.27e+00 | 8.41e-01 | 1.92e+00 | 2.55e-01 |
| Arthralgia(1):Myalgia(1) | 9.98e-01 | 6.67e-01 | 1.49e+00 | 9.92e-01 |
| Arthralgia(1):Food_refusal(1) | 1.36e+00 | 7.20e-01 | 2.56e+00 | 3.44e-01 |
| Arthralgia(1):Tachypnea(1) | 1.29e+00 | 7.57e-01 | 2.20e+00 | 3.49e-01 |
| Arthralgia(1):UAMB(1) | 1.04e+00 | 3.65e-01 | 2.94e+00 | 9.47e-01 |
| Arthralgia(1):Cough(1) | 7.92e-01 | 5.21e-01 | 1.20e+00 | 2.74e-01 |
| Arthralgia(1):Vomiting(1) | 1.87e+00 | 7.74e-01 | 4.54e+00 | 1.64e-01 |
| Headache(1):Mental_confusion(1) | 1.33e+00 | 2.80e-01 | 6.36e+00 | 7.17e-01 |
| Headache(1):Seizures(1) | 3.38e+05 | 1.56e-148 | 7.35e+158 | 9.44e-01 |
| Headache(1):Diarrhea(1) | 1.23e+00 | 7.42e-01 | 2.04e+00 | 4.21e-01 |

| | | | | |
|---|---|---|---|---|
| Headache(1):Dysgeusia(1) | 1.01e+00 | 4.37e-01 | 2.35e+00 | 9.75e-01 |
| Headache(1):Dyspnea(1) | 1.03e+00 | 6.39e-01 | 1.65e+00 | 9.11e-01 |
| Headache(1):Abdominal_pain(1) | 1.61e+00 | 8.74e-01 | 2.96e+00 | 1.26e-01 |
| Headache(1):Chest_pain(1) | 6.74e-01 | 3.57e-01 | 1.27e+00 | 2.23e-01 |
| Headache(1):Low_fever(1) | 1.90e+00 | 9.89e-01 | 3.64e+00 | 5.39e-02 |
| Headache(1):High_fever(1) | 7.39e-01 | 5.21e-01 | 1.05e+00 | 9.00e-02 |
| Headache(1):Respiratory_failure(1) | 7.96e-01 | 4.07e-01 | 1.56e+00 | 5.06e-01 |
| Headache(1):Conjunctival_injection(1) | 5.32e-01 | 2.12e-01 | 1.34e+00 | 1.79e-01 |
| Headache(1):Irritability(1) | 5.19e-01 | 1.20e-01 | 2.24e+00 | 3.80e-01 |
| Headache(1):Malaise(1) | 1.09e+00 | 7.87e-01 | 1.52e+00 | 5.99e-01 |
| Headache(1):Myalgia(1) | 6.63e-01 | 4.53e-01 | 9.68e-01 | 3.36e-02 |
| Headache(1):Food_refusal(1) | 6.24e-01 | 3.40e-01 | 1.15e+00 | 1.28e-01 |
| Headache(1):Tachypnea(1) | 1.02e+00 | 6.40e-01 | 1.63e+00 | 9.31e-01 |
| Headache(1):UAMB(1) | 1.40e+00 | 5.31e-01 | 3.70e+00 | 4.96e-01 |
| Headache(1):Cough(1) | 1.34e+00 | 9.64e-01 | 1.85e+00 | 8.20e-02 |
| Headache(1):Vomiting(1) | 1.20e+00 | 5.60e-01 | 2.58e+00 | 6.35e-01 |
| Mental_confusion(1):Seizures(1) | 5.50e+14 | 0.00e+00 | Inf | 9.39e-01 |
| Mental_confusion(1):Diarrhea(1) | 2.42e+00 | 6.22e-01 | 9.44e+00 | 2.02e-01 |
| Mental_confusion(1):Dysgeusia(1) | 4.86e-07 | 0.00e+00 | Inf | 9.95e-01 |
| Mental_confusion(1):Dyspnea(1) | 4.82e-01 | 1.74e-01 | 1.33e+00 | 1.60e-01 |
| Mental_confusion(1):Abdominal_pain(1) | 1.19e+00 | 2.55e-01 | 5.54e+00 | 8.25e-01 |
| Mental_confusion(1):Chest_pain(1) | 1.68e+00 | 3.24e-01 | 8.74e+00 | 5.36e-01 |
| Mental_confusion(1):Low_fever(1) | 2.59e-01 | 2.41e-02 | 2.79e+00 | 2.65e-01 |
| Mental_confusion(1):High_fever(1) | 8.52e-01 | 3.48e-01 | 2.09e+00 | 7.26e-01 |
| Mental_confusion(1):Respiratory_failure(1) | 2.35e+00 | 8.53e-01 | 6.47e+00 | 9.84e-02 |
| Mental_confusion(1):Conjunctival_injection(1) | 1.87e-07 | 0.00e+00 | Inf | 9.85e-01 |
| Mental_confusion(1):Irritability(1) | 2.11e+00 | 7.82e-01 | 5.68e+00 | 1.40e-01 |
| Mental_confusion(1):Malaise(1) | 4.47e-01 | 1.73e-01 | 1.15e+00 | 9.58e-02 |
| Mental_confusion(1):Myalgia(1) | 1.06e+00 | 2.20e-01 | 5.08e+00 | 9.44e-01 |
| Mental_confusion(1):Food_refusal(1) | 1.60e+00 | 5.47e-01 | 4.65e+00 | 3.92e-01 |
| Mental_confusion(1):Tachypnea(1) | 5.00e+00 | 1.87e+00 | 1.34e+01 | 1.38e-03 |
| Mental_confusion(1):UAMB(1) | 7.73e-01 | 2.39e-01 | 2.50e+00 | 6.68e-01 |
| Mental_confusion(1):Cough(1) | 1.08e+00 | 4.56e-01 | 2.55e+00 | 8.63e-01 |
| Mental_confusion(1):Vomiting(1) | 1.36e+00 | 2.38e-01 | 7.73e+00 | 7.30e-01 |
| Seizures(1):Diarrhea(1) | 3.66e+04 | 0.00e+00 | Inf | 9.92e-01 |
| Seizures(1):Dysgeusia(1) | 6.51e+11 | 0.00e+00 | Inf | 9.92e-01 |
| Seizures(1):Dyspnea(1) | 6.62e+00 | 1.01e-01 | 4.33e+02 | 3.75e-01 |
| Seizures(1):Abdominal_pain(1) | 1.88e-03 | 0.00e+00 | Inf | 9.97e-01 |
| Seizures(1):Chest_pain(1) | 4.35e+14 | 0.00e+00 | Inf | 9.39e-01 |
| Seizures(1):Low_fever(1) | 7.56e-09 | 0.00e+00 | Inf | 9.94e-01 |
| Seizures(1):High_fever(1) | 1.54e+04 | 7.27e-150 | 3.26e+157 | 9.57e-01 |
| Seizures(1):Respiratory_failure(1) | 5.77e-05 | 2.65e-158 | 1.26e+149 | 9.57e-01 |
| Seizures(1):Conjunctival_injection(1) | 5.65e+19 | 0.00e+00 | Inf | 9.39e-01 |
| Seizures(1):Irritability(1) | 3.79e-09 | 6.56e-276 | 2.19e+258 | 9.51e-01 |
| Seizures(1):Malaise(1) | 1.93e-14 | 0.00e+00 | Inf | 9.75e-01 |
| Seizures(1):Myalgia(1) | 3.98e+05 | 1.76e-148 | 9.01e+158 | 9.43e-01 |
| Seizures(1):Food_refusal(1) | 6.67e-06 | 0.00e+00 | Inf | 9.88e-01 |
| Seizures(1):Tachypnea(1) | 8.36e-07 | 0.00e+00 | Inf | 9.86e-01 |
| Seizures(1):UAMB(1) | 5.56e-21 | 0.00e+00 | Inf | 9.60e-01 |

| | | | | |
|---|---|---|---|---|
| Seizures(1):Cough(1) | 2.83e+09 | 5.02e-258 | 1.60e+276 | 9.45e-01 |
| Seizures(1):Vomiting(1) | 2.02e-05 | 0.00e+00 | Inf | 9.90e-01 |
| Diarrhea(1):Dysgeusia(1) | 2.02e+00 | 4.14e-01 | 9.89e+00 | 3.84e-01 |
| Diarrhea(1):Dyspnea(1) | 1.04e+00 | 5.50e-01 | 1.96e+00 | 9.09e-01 |
| Diarrhea(1):Abdominal_pain(1) | 6.20e-01 | 3.28e-01 | 1.17e+00 | 1.42e-01 |
| Diarrhea(1):Chest_pain(1) | 1.23e+00 | 5.36e-01 | 2.81e+00 | 6.27e-01 |
| Diarrhea(1):Low_fever(1) | 6.14e-01 | 1.57e-01 | 2.41e+00 | 4.84e-01 |
| Diarrhea(1):High_fever(1) | 1.40e+00 | 8.37e-01 | 2.34e+00 | 1.99e-01 |
| Diarrhea(1):Respiratory_failure(1) | 4.26e-01 | 1.59e-01 | 1.14e+00 | 8.90e-02 |
| Diarrhea(1):Conjunctival_injection(1) | 1.09e+00 | 3.05e-01 | 3.86e+00 | 8.98e-01 |
| Diarrhea(1):Irritability(1) | 1.42e+00 | 4.05e-01 | 4.96e+00 | 5.86e-01 |
| Diarrhea(1):Malaise(1) | 1.12e+00 | 7.05e-01 | 1.78e+00 | 6.27e-01 |
| Diarrhea(1):Myalgia(1) | 1.30e+00 | 7.68e-01 | 2.20e+00 | 3.29e-01 |
| Diarrhea(1):Food_refusal(1) | 1.93e+00 | 1.04e+00 | 3.57e+00 | 3.70e-02 |
| Diarrhea(1):Tachypnea(1) | 8.90e-01 | 5.01e-01 | 1.58e+00 | 6.90e-01 |
| Diarrhea(1):UAMB(1) | 1.07e+00 | 3.45e-01 | 3.31e+00 | 9.08e-01 |
| Diarrhea(1):Cough(1) | 1.08e+00 | 6.84e-01 | 1.69e+00 | 7.49e-01 |
| Diarrhea(1):Vomiting(1) | 1.12e+00 | 5.25e-01 | 2.37e+00 | 7.76e-01 |
| Dysgeusia(1):Dyspnea(1) | 1.89e+00 | 5.26e-01 | 6.77e+00 | 3.30e-01 |
| Dysgeusia(1):Abdominal_pain(1) | 1.91e+00 | 2.30e-01 | 1.59e+01 | 5.49e-01 |
| Dysgeusia(1):Chest_pain(1) | 2.28e+00 | 3.39e-01 | 1.54e+01 | 3.96e-01 |
| Dysgeusia(1):Low_fever(1) | 6.19e-01 | 1.18e-01 | 3.24e+00 | 5.70e-01 |
| Dysgeusia(1):High_fever(1) | 6.74e-01 | 3.03e-01 | 1.50e+00 | 3.34e-01 |
| Dysgeusia(1):Respiratory_failure(1) | 7.62e-01 | 1.79e-01 | 3.24e+00 | 7.12e-01 |
| Dysgeusia(1):Conjunctival_injection(1) | 1.12e+00 | 7.12e-02 | 1.78e+01 | 9.34e-01 |
| Dysgeusia(1):Irritability(1) | 1.29e+01 | 3.38e-01 | 4.94e+02 | 1.69e-01 |
| Dysgeusia(1):Malaise(1) | 2.77e-01 | 9.13e-02 | 8.40e-01 | 2.33e-02 |
| Dysgeusia(1):Myalgia(1) | 8.06e-01 | 2.16e-01 | 3.01e+00 | 7.48e-01 |
| Dysgeusia(1):Food_refusal(1) | 2.45e+00 | 5.40e-01 | 1.12e+01 | 2.45e-01 |
| Dysgeusia(1):Tachypnea(1) | 6.90e-01 | 1.95e-01 | 2.44e+00 | 5.64e-01 |
| Dysgeusia(1):UAMB(1) | 1.72e-07 | 0.00e+00 | Inf | 9.95e-01 |
| Dysgeusia(1):Cough(1) | 1.02e+00 | 4.68e-01 | 2.24e+00 | 9.53e-01 |
| Dysgeusia(1):Vomiting(1) | 2.45e+00 | 1.12e-01 | 5.36e+01 | 5.69e-01 |
| Dyspnea(1):Abdominal_pain(1) | 8.34e-01 | 4.34e-01 | 1.60e+00 | 5.84e-01 |
| Dyspnea(1):Chest_pain(1) | 9.65e-01 | 5.59e-01 | 1.66e+00 | 8.97e-01 |
| Dyspnea(1):Low_fever(1) | 1.21e+00 | 5.95e-01 | 2.45e+00 | 6.03e-01 |
| Dyspnea(1):High_fever(1) | 1.11e+00 | 7.99e-01 | 1.53e+00 | 5.40e-01 |
| Dyspnea(1):Respiratory_failure(1) | 1.18e+00 | 7.94e-01 | 1.77e+00 | 4.06e-01 |
| Dyspnea(1):Conjunctival_injection(1) | 1.25e+00 | 4.43e-01 | 3.52e+00 | 6.74e-01 |
| Dyspnea(1):Irritability(1) | 5.32e-01 | 2.00e-01 | 1.41e+00 | 2.06e-01 |
| Dyspnea(1):Malaise(1) | 1.05e+00 | 7.48e-01 | 1.47e+00 | 7.80e-01 |
| Dyspnea(1):Myalgia(1) | 1.00e+00 | 6.13e-01 | 1.64e+00 | 9.94e-01 |
| Dyspnea(1):Food_refusal(1) | 1.37e+00 | 7.74e-01 | 2.43e+00 | 2.79e-01 |
| Dyspnea(1):Tachypnea(1) | 1.21e+00 | 8.84e-01 | 1.65e+00 | 2.36e-01 |
| Dyspnea(1):UAMB(1) | 1.30e+00 | 7.49e-01 | 2.26e+00 | 3.51e-01 |
| Dyspnea(1):Cough(1) | 9.95e-01 | 7.22e-01 | 1.37e+00 | 9.75e-01 |
| Dyspnea(1):Vomiting(1) | 1.16e+00 | 4.73e-01 | 2.84e+00 | 7.47e-01 |
| Abdominal_pain(1):Chest_pain(1) | 2.68e+00 | 1.23e+00 | 5.87e+00 | 1.35e-02 |
| Abdominal_pain(1):Low_fever(1) | 5.32e-01 | 1.53e-01 | 1.85e+00 | 3.21e-01 |

| | | | | |
|---|---|---|---|---|
| Abdominal_pain(1):High_fever(1) | 5.60e-01 | 3.34e-01 | 9.39e-01 | 2.78e-02 |
| Abdominal_pain(1):Respiratory_failure(1) | 9.15e-01 | 4.04e-01 | 2.07e+00 | 8.30e-01 |
| Abdominal_pain(1):Conjunctival_injection(1) | 1.57e+00 | 4.51e-01 | 5.49e+00 | 4.77e-01 |
| Abdominal_pain(1):Irritability(1) | 6.49e-01 | 1.44e-01 | 2.92e+00 | 5.74e-01 |
| Abdominal_pain(1):Malaise(1) | 5.51e-01 | 3.22e-01 | 9.42e-01 | 2.94e-02 |
| Abdominal_pain(1):Myalgia(1) | 9.13e-01 | 4.56e-01 | 1.83e+00 | 7.97e-01 |
| Abdominal_pain(1):Food_refusal(1) | 1.02e+00 | 4.88e-01 | 2.11e+00 | 9.67e-01 |
| Abdominal_pain(1):Tachypnea(1) | 1.69e+00 | 9.08e-01 | 3.16e+00 | 9.79e-02 |
| Abdominal_pain(1):UAMB(1) | 1.27e+00 | 3.82e-01 | 4.20e+00 | 6.99e-01 |
| Abdominal_pain(1):Cough(1) | 1.09e+00 | 6.57e-01 | 1.81e+00 | 7.38e-01 |
| Abdominal_pain(1):Vomiting(1) | 1.77e+00 | 8.27e-01 | 3.81e+00 | 1.41e-01 |
| Chest_pain(1):Low_fever(1) | 3.17e-01 | 3.87e-02 | 2.59e+00 | 2.83e-01 |
| Chest_pain(1):High_fever(1) | 1.77e+00 | 1.07e+00 | 2.90e+00 | 2.48e-02 |
| Chest_pain(1):Respiratory_failure(1) | 1.16e+00 | 6.03e-01 | 2.21e+00 | 6.64e-01 |
| Chest_pain(1):Conjunctival_injection(1) | 1.59e+00 | 5.48e-01 | 4.61e+00 | 3.94e-01 |
| Chest_pain(1):Irritability(1) | 9.01e-01 | 1.86e-01 | 4.37e+00 | 8.97e-01 |
| Chest_pain(1):Malaise(1) | 1.23e+00 | 7.46e-01 | 2.01e+00 | 4.23e-01 |
| Chest_pain(1):Myalgia(1) | 6.13e-01 | 3.26e-01 | 1.15e+00 | 1.29e-01 |
| Chest_pain(1):Food_refusal(1) | 1.02e+00 | 4.54e-01 | 2.30e+00 | 9.59e-01 |
| Chest_pain(1):Tachypnea(1) | 1.27e+00 | 7.54e-01 | 2.15e+00 | 3.66e-01 |
| Chest_pain(1):UAMB(1) | 1.02e+00 | 4.23e-01 | 2.46e+00 | 9.66e-01 |
| Chest_pain(1):Cough(1) | 9.45e-01 | 5.76e-01 | 1.55e+00 | 8.23e-01 |
| Chest_pain(1):Vomiting(1) | 9.82e-02 | 1.14e-02 | 8.46e-01 | 3.47e-02 |
| Low_fever(1):Respiratory_failure(1) | 7.88e-01 | 2.88e-01 | 2.16e+00 | 6.44e-01 |
| Low_fever(1):Conjunctival_injection(1) | 1.56e+00 | 2.19e-01 | 1.11e+01 | 6.56e-01 |
| Low_fever(1):Irritability(1) | 7.96e+00 | 1.48e+00 | 4.28e+01 | 1.56e-02 |
| Low_fever(1):Malaise(1) | 4.91e-01 | 2.62e-01 | 9.18e-01 | 2.58e-02 |
| Low_fever(1):Myalgia(1) | 3.45e-01 | 1.27e-01 | 9.43e-01 | 3.80e-02 |
| Low_fever(1):Food_refusal(1) | 2.59e+00 | 7.65e-01 | 8.77e+00 | 1.26e-01 |
| Low_fever(1):Tachypnea(1) | 1.06e+00 | 5.04e-01 | 2.25e+00 | 8.70e-01 |
| Low_fever(1):UAMB(1) | 7.58e-01 | 1.83e-01 | 3.15e+00 | 7.03e-01 |
| Low_fever(1):Cough(1) | 1.15e+00 | 6.90e-01 | 1.93e+00 | 5.87e-01 |
| Low_fever(1):Vomiting(1) | 2.04e+00 | 4.34e-01 | 9.60e+00 | 3.66e-01 |
| High_fever(1):Respiratory_failure(1) | 1.73e+00 | 1.17e+00 | 2.57e+00 | 6.30e-03 |
| High_fever(1):Conjunctival_injection(1) | 8.40e-01 | 3.56e-01 | 1.98e+00 | 6.91e-01 |
| High_fever(1):Irritability(1) | 1.31e+00 | 5.58e-01 | 3.09e+00 | 5.32e-01 |
| High_fever(1):Malaise(1) | 6.80e-01 | 5.15e-01 | 8.98e-01 | 6.47e-03 |
| High_fever(1):Myalgia(1) | 7.71e-01 | 5.27e-01 | 1.13e+00 | 1.80e-01 |
| High_fever(1):Food_refusal(1) | 1.34e+00 | 7.64e-01 | 2.35e+00 | 3.07e-01 |
| High_fever(1):Tachypnea(1) | 1.17e+00 | 8.52e-01 | 1.60e+00 | 3.33e-01 |
| High_fever(1):UAMB(1) | 7.38e-01 | 4.22e-01 | 1.29e+00 | 2.85e-01 |
| High_fever(1):Cough(1) | 1.16e+00 | 8.96e-01 | 1.49e+00 | 2.63e-01 |
| High_fever(1):Vomiting(1) | 2.13e+00 | 8.68e-01 | 5.21e+00 | 9.88e-02 |
| Respiratory_failure(1):Conjunctival_injection(1) | 7.41e-01 | 2.05e-01 | 2.68e+00 | 6.47e-01 |
| Respiratory_failure(1):Irritability(1) | 3.49e-01 | 1.20e-01 | 1.02e+00 | 5.38e-02 |
| Respiratory_failure(1):Malaise(1) | 1.06e+00 | 7.03e-01 | 1.60e+00 | 7.79e-01 |
| Respiratory_failure(1):Myalgia(1) | 8.26e-01 | 4.25e-01 | 1.61e+00 | 5.75e-01 |
| Respiratory_failure(1):Food_refusal(1) | 7.68e-01 | 3.72e-01 | 1.59e+00 | 4.76e-01 |
| Respiratory_failure(1):Tachypnea(1) | 1.33e+00 | 8.96e-01 | 1.96e+00 | 1.59e-01 |

| | | | | |
|---|---|---|---|---|
| Respiratory_failure(1):UAMB(1) | 1.19e+00 | 6.78e-01 | 2.08e+00 | 5.47e-01 |
| Respiratory_failure(1):Cough(1) | 1.24e+00 | 8.50e-01 | 1.82e+00 | 2.61e-01 |
| Respiratory_failure(1):Vomiting(1) | 1.90e+00 | 6.54e-01 | 5.51e+00 | 2.39e-01 |
| Conjunctival_injection(1):Irritability(1) | 7.62e-01 | 9.33e-02 | 6.23e+00 | 8.00e-01 |
| Conjunctival_injection(1):Malaise(1) | 1.23e+00 | 5.46e-01 | 2.78e+00 | 6.15e-01 |
| Conjunctival_injection(1):Myalgia(1) | 1.87e+00 | 7.54e-01 | 4.65e+00 | 1.76e-01 |
| Conjunctival_injection(1):Food_refusal(1) | 1.14e+00 | 3.26e-01 | 3.97e+00 | 8.40e-01 |
| Conjunctival_injection(1):Tachypnea(1) | 1.04e+00 | 3.89e-01 | 2.79e+00 | 9.36e-01 |
| Conjunctival_injection(1):UAMB(1) | 1.33e+00 | 3.00e-01 | 5.93e+00 | 7.06e-01 |
| Conjunctival_injection(1):Cough(1) | 5.53e-01 | 2.51e-01 | 1.22e+00 | 1.43e-01 |
| Conjunctival_injection(1):Vomiting(1) | 1.53e-07 | 0.00e+00 | Inf | 9.86e-01 |
| Irritability(1):Malaise(1) | 2.15e+00 | 9.35e-01 | 4.95e+00 | 7.14e-02 |
| Irritability(1):Myalgia(1) | 9.03e-01 | 2.32e-01 | 3.52e+00 | 8.83e-01 |
| Irritability(1):Food_refusal(1) | 1.20e+00 | 4.72e-01 | 3.04e+00 | 7.05e-01 |
| Irritability(1):Tachypnea(1) | 5.13e-01 | 2.12e-01 | 1.24e+00 | 1.38e-01 |
| Irritability(1):UAMB(1) | 5.13e+00 | 1.75e+00 | 1.51e+01 | 2.95e-03 |
| Irritability(1):Cough(1) | 5.97e-01 | 2.71e-01 | 1.32e+00 | 2.02e-01 |
| Irritability(1):Vomiting(1) | 9.08e-01 | 1.72e-01 | 4.80e+00 | 9.10e-01 |
| Malaise(1):Myalgia(1) | 8.69e-01 | 6.13e-01 | 1.23e+00 | 4.29e-01 |
| Malaise(1):Food_refusal(1) | 9.89e-01 | 6.02e-01 | 1.63e+00 | 9.65e-01 |
| Malaise(1):Tachypnea(1) | 1.19e+00 | 8.55e-01 | 1.65e+00 | 3.06e-01 |
| Malaise(1):UAMB(1) | 8.69e-01 | 4.82e-01 | 1.57e+00 | 6.40e-01 |
| Malaise(1):Cough(1) | 8.52e-01 | 6.55e-01 | 1.11e+00 | 2.33e-01 |
| Malaise(1):Vomiting(1) | 9.73e-01 | 4.88e-01 | 1.94e+00 | 9.37e-01 |
| Myalgia(1):Food_refusal(1) | 1.30e+00 | 7.33e-01 | 2.29e+00 | 3.73e-01 |
| Myalgia(1):Tachypnea(1) | 8.82e-01 | 5.45e-01 | 1.43e+00 | 6.11e-01 |
| Myalgia(1):UAMB(1) | 1.42e+00 | 5.67e-01 | 3.54e+00 | 4.56e-01 |
| Myalgia(1):Cough(1) | 1.29e+00 | 8.98e-01 | 1.86e+00 | 1.68e-01 |
| Myalgia(1):Vomiting(1) | 9.39e-01 | 4.17e-01 | 2.11e+00 | 8.80e-01 |
| Food_refusal(1):Tachypnea(1) | 1.30e+00 | 7.47e-01 | 2.26e+00 | 3.53e-01 |
| Food_refusal(1):UAMB(1) | 3.53e-01 | 1.22e-01 | 1.02e+00 | 5.43e-02 |
| Food_refusal(1):Cough(1) | 1.35e+00 | 8.09e-01 | 2.26e+00 | 2.49e-01 |
| Food_refusal(1):Vomiting(1) | 2.39e+00 | 1.09e+00 | 5.26e+00 | 2.99e-02 |
| Tachypnea(1):UAMB(1) | 8.79e-01 | 4.90e-01 | 1.58e+00 | 6.65e-01 |
| Tachypnea(1):Cough(1) | 7.49e-01 | 5.49e-01 | 1.02e+00 | 6.74e-02 |
| Tachypnea(1):Vomiting(1) | 7.79e-01 | 3.38e-01 | 1.80e+00 | 5.59e-01 |
| UAMB(1):Cough(1) | 9.76e-01 | 5.43e-01 | 1.76e+00 | 9.35e-01 |
| UAMB(1):Vomiting(1) | 1.88e+00 | 4.97e-01 | 7.11e+00 | 3.52e-01 |
| Cough(1):Vomiting(1) | 6.93e-01 | 3.55e-01 | 1.35e+00 | 2.82e-01 |
| Sex(M):Odynophagia(1) | 1.87e+00 | 9.20e-01 | 3.82e+00 | 8.37e-02 |
| Anosmia(1):Odynophagia(1) | 7.72e+00 | 9.21e-01 | 6.46e+01 | 5.95e-02 |
| Arthralgia(1):Odynophagia(1) | 8.07e-01 | 1.79e-01 | 3.64e+00 | 7.80e-01 |
| Headache(1):Odynophagia(1) | 9.66e-01 | 4.11e-01 | 2.27e+00 | 9.36e-01 |
| Mental_confusion(1):Odynophagia(1) | 2.00e-41 | 0.00e+00 | Inf | 9.99e-01 |
| Diarrhea(1):Odynophagia(1) | 3.24e+00 | 2.17e-01 | 4.83e+01 | 3.94e-01 |
| Dysgeusia(1):Odynophagia(1) | 1.01e-01 | 1.38e-02 | 7.46e-01 | 2.46e-02 |
| Dyspnea(1):Odynophagia(1) | 2.17e+00 | 6.70e-01 | 7.04e+00 | 1.96e-01 |
| Abdominal_pain(1):Odynophagia(1) | 4.84e+00 | 3.84e-01 | 6.10e+01 | 2.22e-01 |
| Chest_pain(1):Odynophagia(1) | 1.40e+00 | 2.27e-01 | 8.57e+00 | 7.19e-01 |

| | | | | |
|---|---|---|---|---|
| Low_fever(1):Odynophagia(1) | 9.03e-01 | 2.66e-01 | 3.07e+00 | 8.70e-01 |
| High_fever(1):Odynophagia(1) | 2.44e+00 | 1.12e+00 | 5.29e+00 | 2.42e-02 |
| Respiratory_failure(1):Odynophagia(1) | 9.26e-01 | 1.85e-01 | 4.63e+00 | 9.26e-01 |
| Conjunctival_injection(1):Odynophagia(1) | 9.10e+19 | 0.00e+00 | Inf | 9.99e-01 |
| Irritability(1):Odynophagia(1) | 2.09e+77 | 0.00e+00 | Inf | 9.99e-01 |
| Malaise(1):Odynophagia(1) | 1.12e+00 | 4.73e-01 | 2.64e+00 | 8.01e-01 |
| Myalgia(1):Odynophagia(1) | 1.84e+00 | 5.34e-01 | 6.36e+00 | 3.33e-01 |
| Odynophagia(1):Food_refusal(1) | 1.14e-40 | 0.00e+00 | Inf | 9.97e-01 |
| Odynophagia(1):Tachypnea(1) | 9.87e-01 | 2.84e-01 | 3.43e+00 | 9.83e-01 |
| Odynophagia(1):UAMB(1) | 1.14e+00 | 7.66e-02 | 1.70e+01 | 9.23e-01 |
| Odynophagia(1):Cough(1) | 9.13e-01 | 4.43e-01 | 1.88e+00 | 8.05e-01 |
| Odynophagia(1):Vomiting(1) | 4.86e+00 | 0.00e+00 | Inf | 9.99e-01 |
