## Supplementary material for "Understanding the value of clinical symptoms of COVID-19. A logistic regression model": S4 Appendix.pdf

**S4 Appendix. Alternative model with the variables sex, age, 20 symptoms and pairwise interactions.**

This alternative model has an AIC = 34986 and an AUC = 0.7127, with a p = 0.08436 it has a sensitivity and specificity of 0.80(0.70,0.82). The equation is described below, where p is the probability that the RT-PCR for SARS-CoV-2 will be positive, symptom present = 1, symptom absent = 0, male = 1 and female = 0.

$$p = \frac{1}{(1 + e^{-B})}$$

B= -1.626354-0.013390*age+0.134393*Sex+2.834388*Anosmia-0.051723*Arthralgia+0.7205*Headache-1.047505*Mental confusion-0.806439*Seisures-0.614470*Diarrhea+1.171302*Dysgeusia-0.239434*Dysnea-0.354076*Abdominal pain-0.382208*Chest pain+0.137223*Low fever-0.972277*High fever-0.364329*Conjuctival injection-0.404076*Irritability-0.013226*Malaise+0.212211*Myalgias-0.391906*Food refusal-0.540014*Tachypnea+0.136984*Cough-0.693652*Vomiting-0.017895*age*Anosmia-0.004823*age*Headache+0.008238*age*diarrea+0.009578*age*low grade fever+0.017499*age*High grade fever+0.002822*age*Cough-0.746708*Anosmia*Dysgeusia+0.968035*Anosmia*Conjuctival injection-0.504919*Anosmia*Malaise-0.269136*Arthralgia*Headache+0.602225*artralgia*Chest pain+0.492844*Arthralgia*Food refusal-0.255732*Headache*Myalgias+0.861888*Mental confusion*Irritability+1.980105*Seisures*Diarrhea-1.280320*Seisures*High fever+0.563359*Diarrhea*Chest pain+0.831252*Disgeusia*Tachypnea+0.693108*Dyspnea*Conjunctival injection-0.297839*Abdominal pain*High grade fever+0.541249*Abdominal pain*Tachypnea+0.512155*Abdominal pain*Vomiting-0.305776*Chest pain*Myalgia-0.182407*High grade fever*Malaise+ 0.249264*High grade fever*Tachypnea+0.219189*High grade fever*Cough-0.265721*Tachypnea*Cough
